# Plantar Thermography for Longitudinal Remote Monitoring of Chronic Heart Failure

**DOI:** 10.64898/2026.10.20.26364428

**Authors:** Michele L. Esposito, Lefan Xuan, Elizabeth Zemp, Michael R. Zile, Ryan J. Tedford, David R. Linders, Kurt W. Prins

## Abstract

Rural patients with heart failure (HF) have poor access to care, worse outcomes, and may benefit from at-home monitoring. We evaluated whether foot temperature measured using a thermographic floor mat was associated with invasively measured cardiac output and whether longitudinal temperature changes preceded HF events. We conducted a two-part prospective, observational single-center study using the SmartMat (Podimetrics, Somerville, MA), which acquires plantar thermograms and ambient temperature. In the acute study, patients undergoing clinically indicated right heart catheterization completed 4 scans on the same day. In the chronic study, patients with a HF hospitalization in the past year and New York Heart Association Class II-III symptoms were instructed to use the device and a scale daily for 6 months. A HF event was defined as receiving at least one dose of intravenous or subcutaneous diuretic. Thirty patients were enrolled in the acute study. Mean right hallux temperature positively correlated with thermodilution cardiac output (r=0.63, p<0.001). 42 patients were enrolled in the chronic study. Among patients experiencing an HF event, ambient-adjusted hallux temperature declined in the 15 days immediately preceding the event compared with days 15– 30 before the event. Plantar thermograms are associated with invasively measured cardiac output and may detect subclinical worsening before HF events. Foot temperature may provide complementary information regarding HF decompensation, supporting further evaluation of plantar thermography for remote HF monitoring.

---

Heart failure (HF) is a leading public health burden for the United States.^1^ At present, there are limited at-home, non-invasive device monitoring programs that can detect subtle HF decompensation, which is a particular concern for patients with limited heart care touch-points. In the US, rural patients with HF suffer from poor access to care, worse clinical outcomes, and higher rates of emergency/acute care visits than their non-rural counterparts. Nationally, 46% of U.S. counties have no practicing cardiologist, which increases to 86% for rural counties, leaving approximately 22 million Americans without access to a cardiologist in their county.^2^ Additionally, patients living in those rural counties need to travel for an average 87 miles roundtrip to even meet with a cardiology provider.^2^ There is therefore a large, underexplored opportunity to increase telemedical care by deploying low-cost monitoring systems into these underserved communities.

Retrospective studies have demonstrated an association between decreasing body temperature and HF admissions.^3,4^ Additionally, correlation between great toe temperature and cardiac output has been shown.^5^ However, a study measuring lower extremity temperature through longitudinal, home-based thermography has never been performed. We therefore evaluated whether foot temperature measured using a thermographic floor mat was associated with invasively measured cardiac output and whether longitudinal temperature changes preceded HF events.

We conducted a two-part prospective, observational single-center study using the SmartMat (Podimetrics, Somerville, Massachusetts, **Figure 1A**), a connected device that acquires thermograms of the plantar surfaces of both feet, as well as ambient temperature, and transmits this data wirelessly to a HIPAA-compliant server for offline analysis. For each foot, 6 separate temperature datapoints were acquired: hallux, 1^st^ metatarsal head, 3^rd^ metatarsal head, 5^th^ metatarsal head, heel, and medial arch (**Figure 1B**). Both raw temperatures and the difference in temperature minus ambient temperature were recorded. In the acute study, patients undergoing a clinically indicated right heart catheterization (RHC) completed four different scans on the same day as their RHC. Temperature data was averaged over the four scans for each of the 6 anatomic locations. In the chronic study, patients with a HF hospitalization within the preceding year and New York Heart Association (NYHA) Class II-III symptoms, were recruited. Study subjects were sent home with both the study device as well as a weight scale (Withings, Inc) and were instructed to stand on both daily for 6 months. A HF event visit was defined as administration of at least one dose of intravenous or subcutaneous diuretic for the diagnosis of either systolic or diastolic HF. Exclusion criteria for both studies included implantation of an acute or durable mechanical circulatory support device, history of congenital heart disease, active Charcot disease, presence of plantar foot ulcers or gangrene, lower limb amputation, end-stage renal disease requiring dialysis, or active pregnancy.

**Figure 1.**
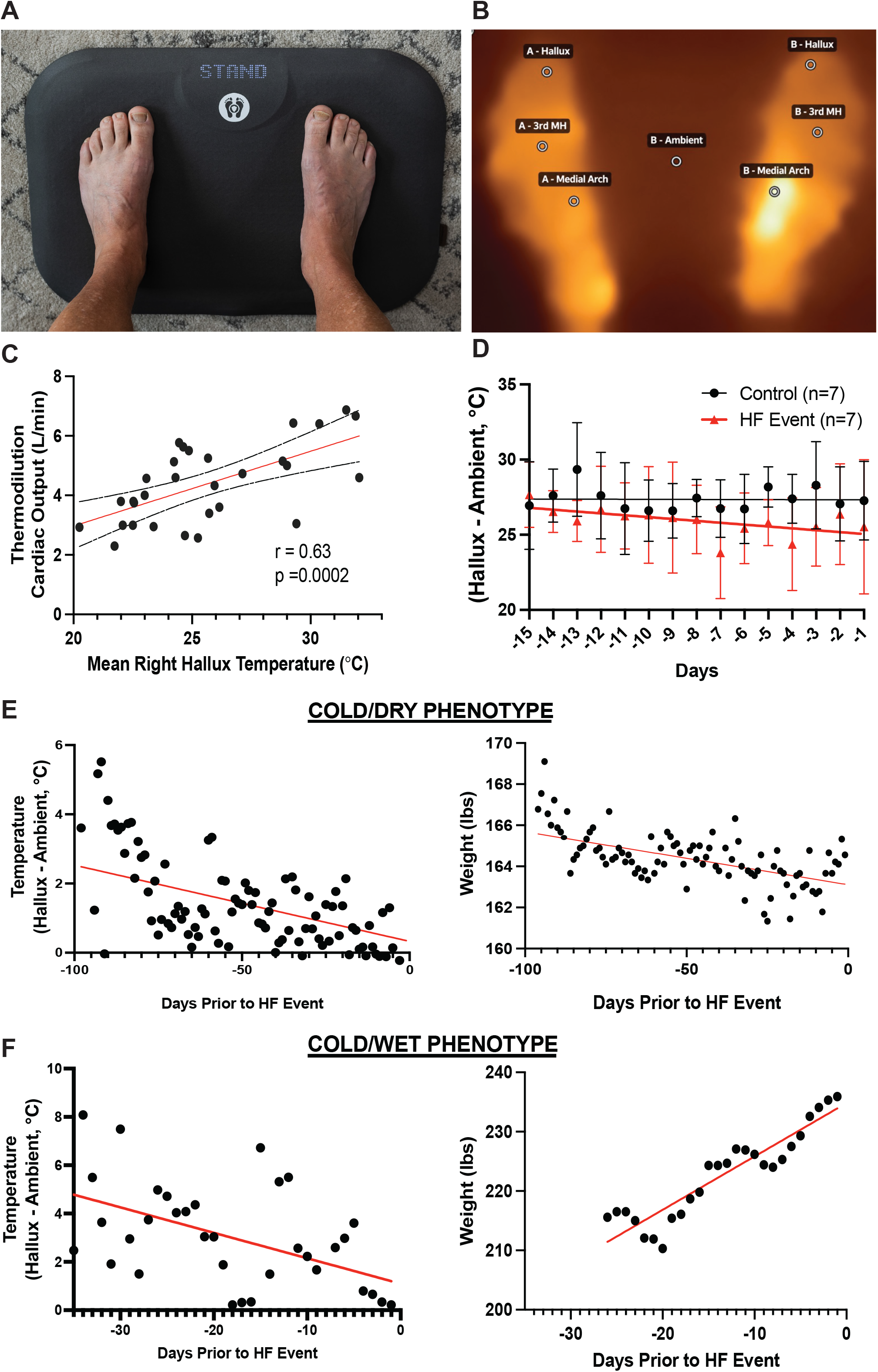
A: SmartMat device (courtesy of Podimetrics, Inc). B: Example plantar thermogram of the soles of both feet. C: Association between temperature of the right hallux and thermodilution cardiac output among the total cohort of the acute study. D: Aggregate patient data showing mean temperatures (minimum hallux temperature– ambient temperature) with standard error bars over time. Study subjects serve as their own controls. Black dots represent temperature from 15-30 days prior to a HF event (where Day=-15 signifies 30 days out from a hospitalization) and red boxes represent the 15 days prior to a hospitalization where Day = -15 signifies 15 days out from a HF event. E: Weight and foot temperature readings from a patient in his 70s with chronic systolic HF and NYHA Class III symptoms requiring admission for HF exacerbation. F: Recordings of weight and foot temperature from a patient in his 50s with chronic systolic HF and NYHA Class III prior to admission for HF exacerbation.

In the acute study, 30 patients were enrolled. Mean age was 52±13 years and left ventricular ejection fraction (LVEF) was 35±20%. Average thermodilution cardiac output and index were 4.4±1.3 L/min and 2.2±0.7 L/min/m^2^ respectively. There was a significant positive correlation between mean right hallux temperature and thermodilution cardiac output (*r*=0.63, p=0.0002) (**Figure 1C**). In the longitudinal phase study, 42 patients were enrolled, of which 31 (74%) were male. Mean age and LVEF was 66±13 years and 30±13%. Of the total cohort, 15 (36%) had an ischemic HF etiology. Only 3 (7%) patients had HF with preserved EF, defined as EF ≥ 50%. Over the study period, there were 17 HF events among 13 patients (31%). Among patients with at least one HF event and both temperature and weight data available from 30 days prior to the event, there was a downtrend in hallux temperature during the 15 days preceding the event, compared to days 15–30 before the event, which served as control. (**Figure 1D**). Longitudinal temperature and weight data from two patients with NYHA Class III chronic systolic HF (**Figure 1E-F)** are shown, illustrating how temperature data might provide complementary information to weight alone. **Figure 1E** shows a patient with a reduction in hallux temperature and mild weight loss about 100 days prior to a HF hospitalization, consistent with a cold/dry phenotype, whereas **Figure 1F** reveals a patient with both weight gain and reduction in hallux temperature approximately 30 days prior to a HF hospitalization, consistent with a cold/wet phenotype.

In this proof-of-concept study, we show that plantar thermograms are associated with invasively-derived cardiac output and are able to longitudinally detect subclinical worsening prior to a HF event. Unlike weight, which primarily reflects fluid overload, peripheral foot temperature may provide complementary data regarding reduced perfusion, the “cold” phenotype from established HF hemodynamic profiles. These hypothesis-generating findings support further evaluation of plantar thermography as a scalable, noninvasive approach to remote HF monitoring, particularly for vulnerable patients with limited access to subspecialty care.

## Data Availability

All data produced in the present study are available upon reasonable request to the authors

## Acknowledgements

The authors would like to acknowledge Podimetrics, Inc. (Somerville, MA) for providing the SmartMat™ devices and data monitoring used to collect temperature in this study.

## References

1. Martin, S. S. et al. 2025 Heart Disease and Stroke Statistics: A Report of US and Global Data From the American Heart Association. Circulation 151, e41–e660 (2025).

2. Kim, J. H., Cisneros, T., Nguyen, A., van Meijgaard, J. & Warraich, H. J. Geographic Disparities in Access to Cardiologists in the United States. J. Am. Coll. Cardiol. 84, 315–316 (2024).

3. Omar, H.R., Charnigo, R. & Guglin, M. Admission-to-discharge temperature reduction in decompensated heart failure is associated with rehospitalization. Herz 43, 649–655 (2018).

4. Ahmed, A. et al. Decreasing body temperature predicts early rehospitalization in congestive heart failure. J. Card. Fail. 14, 489–496 (2008).

5. Joly, H. R. & Weil, M. H. Temperature of the great toe as an indication of the severity of shock. Circulation 39, 131–138 (1969).

